# Association of circulating Fibroblast Growth Factor-2 with progression of HIV-chronic kidney diseases in children

**DOI:** 10.1101/2020.12.08.20246280

**Authors:** Patricio E Ray, Jinliang Li, Jharna Das, Jing Yu

## Abstract

**Introduction:** People living with HIV frequently show high plasma levels of Fibroblast Growth Factor-2 (FGF-2/bFGF). Previous studies reported that FGF-2 can accelerate the progression of experimental kidney diseases. However, how circulating FGF-2 affects the progression of HIV-chronic kidney diseases (HIV-CKDs) in children is unknown.

**Methods:** To address this question we measured the plasma and urine levels of FGF-2 in 84 children (< 12 years of age) living with HIV, and determine their association with a high viral load (HVL) and HIV-CKDs. Kidney sections from children with HIV-CKD were used to assess the localization and expression levels of the FGF-2 binding sites. The fate of circulating FGF-2 was determined in young wild type and HIV-transgenic (HIV-Tg_26_) mice injected with human recombinant FGF-2. Cells cultured from children with HIV-CKDs where used to define how FGF-2 affected their infection, survival, and expression of APOL1.

**Results:** High plasma FGF-2 levels were associated with a HVL and HIV-CKDs. High urine FGF-2 levels were found in almost all children with HIV-CKDs. A large reservoir of renal FGF-2 low affinity binding sites in children and HIV-Tg_26_ mice with HIV-CKDs facilitated the recruitment of circulating FGF-2. FGF-2 slightly decreased the expression of APOL1 mRNA in cultured podocytes, but increased the survival of HIV infected inflammatory cells or podocytes, and precipitated HIV-nephropathy in HIV-Tg_26_ mice.

**Conclusion:** Children with high plasma and urine FGF-2 levels were more likely to develop HIV-CKDs. Persistently high plasma FGF-2 levels appear to be an independent risk factor for developing progressive childhood HIV-CKDs.

## INTRODUCTION

People of African ancestry living with the Human Immunodeficiency Virus (HIV-1) and not receiving appropriate antiretroviral therapy (ART) are at high risk for developing HIV-associated nephropathy (HIVAN) and other HIV-chronic kidney diseases (HIV-CKDs).^1^ The majority of the 1.7 million children aged 0 −14 living with HIV worldwide are from African ancestry, and therefore are at high risk of developing HIV-CKDs.^2^ Furthermore, providing long-term ART to children until they reach adulthood is a major public health challenge, and we continue to see children and adolescents with HIV-CKDs in the Washington DC area.^3^ Therefore, more work needs to be done to prevent the development of HIV-CKDs in children.

The Apolipoprotein L1 (APOL1) risks alleles found mainly in people of African ancestry are a major risk factor for the development of HIVAN. ^4-6^ People homozygous for the APLO1 G1 or G2 renal risk alleles (or G1/G2 compound heterozygotes) have a 30-fold increased risk of HIVAN. ^7^ However, the role that the APOL1 risk alleles play in the pathogenesis of HIVAN in young children is less clearly understood. In addition, given the relative low prevalence of the APOL1 risk alleles in people from African ancestry, the majority of children with HIV-CKDs do not carry these risk alleles. ^5, 6^ Thus, it is necessary to identify additional risk factors for developing childhood HIV-CKDs.

Previous studies showed that a high viral load is a major risk factor for developing childhood HIV-CKDs.^3 5^ HIV-infected cells release cytokines and inflammatory mediators that may affect the outcome of HIV-CKDs, however, very few studies have explored this hypothesis in children. High plasma levels of Fibroblast Growth Factor-2 (FGF-2 or basic FGF) were detected in children^8^ and adults^9^ living with HIV. FGF-2 is an heparin binding growth factor that affects the outcome of several experimental renal diseases in different animal models^10-15^, including HIV-transgenic (HIV-Tg_26_) mice.^16^ Nonetheless, the role of circulating FGF-2 in the pathogenesis of childhood HIV-CKDs is unclear at the present time. Here, we hypothesized that high levels of circulating FGF-2, sustained for a prolonged period of time, can accelerate the progression of HIV-CKDs in children. Therefore, we carried out this study to define the association between circulating FGF-2 and childhood HIV-CKDs, and to identify potential mechanisms through which FGF-2 may affect the outcome of these renal diseases.

## METHODS

### Collection of plasma, urine, and kidney samples

All plasma and urine samples were collected from children living with HIV (≤ 12 years of age) who received standard of care every 3 months at Children’s National Hospital (CNH) in Washington, DC. between January 1994 and December 2000. Only samples from children who completed at least 3 years of follow up with annual assessments of viral load, proteinuria, and serum creatinine (SCr) were used in this study. Based on clinical criteria available at that time^17^, a high viral load was defined by at least 3 measurements > 5,000 (3.7 log_10_) HIV-RNA copies/ml within the 6 −12 months prior to the FGF-2 measurement. HIV-CKD was defined by the presence of persistent proteinuria, 1+ dipstick or Upr/Cr ratio > 0.2, and/or an elevated SCr level, for at least two consecutive years, and by renal histology whenever available. Frozen or formalin-fixed archival renal biopsies or autopsies were collected from 9 control children without renal diseases, and 9 children with HIV-CKDs (HIVAN = 4, HIV-associated Hemolytic Uremic syndrome =1; and HIV-immune complex kidney diseases = 4). This study was carried out in accordance with the Declaration of Helsinki and approved by the Institutional Review Board (IRB) at CNH.

### FGF-2 measurements

All samples were centrifuged at 4,000 x g for 5 minutes and stored immediately at −80°C until processed. FGF-2 levels were measured with the ELISA Quantikine Kit (R&D Systems, Minneapolis, MN) as previously described.^8^ Inter- and intra-assay coefficient of variation for FGF-2 was 3-9%. FGF-2 urinary levels were expressed as a ratio of the urinary creatinine (UCr) concentration, in pg per mg of UCr. All measurements were made in duplicate and investigators performing assays were blinded to the clinical outcomes.

### FGF-2 binding studies

[^125^I]-FGF-2 binding studies were done by autoradiography as described before.^16, 18^ Briefly, frozen renal sections (16 µm) were preincubated for 15 minutes in binding buffer (Dulbecco’s Modified Eagle’s Medium [DMEM], 20 mM HEPES at pH 7.4, and 0.15% gelatin), and exposed to 2 ng/ml [^125^I] FGF −2 (Amersham Corp., Arlington Heights, IL, USA) at 4°C for two hours. Nonspecific binding was determined by displacing the binding of [^125^I] FGF −2 with 300 µg/ml heparin (Fisher Scientific, New Jersey, USA) as describe before.^16^ All slides were washed, dried, and exposed to an autoradiography film inside a cassette under similar conditions at room temperature for up to four days. Differences in binding were determined by counting the optical density of the autoradiograms in specific kidney areas of 0.22 mm^2^ using a computerized program as described before.^16, 18^

### Renal recruitment of circulating FGF-2

These experiments were conducted in accordance with the US National Institutes of Health *Guide for the Care and Use of Laboratory Animals* and approved by the Animal Care and use Committee at CNH. Three weeks old FVB/N wild type or HIV-Tg_26_ male mice (n = 3 per group) were anesthetized with ketamine and xylazine (70 - 7 mg/kg/body weight) and injected intravenously with 0.1 ng of [^125^I] FGF-2 diluted in 100 µl of normal saline (specific activity 1 µCi/ng). Subsequently 20µl of blood were collected from the left eye by retro-orbital bleeding at different time points, and all mice were euthanized by cervical dislocation under anesthesia to remove the kidneys. The radioactivity of the blood and kidney samples was measured with a gamma counter as previously described.^19^

### Induction of renal injury by human recombinant (hr)FGF-2

Endotoxin free hr-FGF-2 was purchased from R&D Systems. Three weeks old wild type or HIV-Tg_26_ FVB/N male mice (n= 5 per group) were injected intraperitoneally with FGF-2 (10 µg daily for 7 consecutive days), and sacrificed by cervical dislocation under halothane anesthesia. Renal sections were harvested and processed for light and electron-microscopy as described before.^16^ The percentage of glomerular and tubular structures exhibiting segmental or global sclerosis, tubular dilatation/casts, positive PCNA staining, and proteinuria were combined to generate renal injury scores: 0 = 0-5%; 1 = >5 –10%; 2 = >10 - 25%; 3= >25 - 50%; 4 = > 50%.

### Immunhistochemistry and Alcian blue staining

The FGF-2 and PCNA staining was done as described before.^20, 21^ Briefly, renal sections were incubated for 1 h at room temperature with affinity purified IgG fractions (2.5 µg/ml) of a rabbit polyclonal antibody directed against a unique peptide sequence of FGF-2 (provided by Dr. Baird, PRIZM Pharmaceuticals, San Diego CA, USA), or a monoclonal antibody (clone PC10) against the proliferative cell nuclear antigen (PCNA) from Zymed Laboratories (South San Francisco, CA, USA). These antibodies were validated in previous rodent and human studies.^20, 22^ To facilitate penetration of the FGF-2 antibody, sections were treated with 1 mg/ml of hyaluronidase (type V: Sigma) buffered at pH 5.5 with 0.1 M Sodium Acetate containing 0.15 M NaC1, for 30 minutes at 37°C. Nonimmune rabbit IgGs were used for the negative controls as described before.^22, 23^ For the Alcian blue staining, all sections were placed in 0.1N HCl acid for 1 minute, incubated with a 1% solution at pH 1 Alcian blue (ab150661 from Abcam) for 30 minutes, and then washed and stained with nuclear-fast red as described before. The FGF-2 and Alcian blue staining was quantified in five different glomerular and tubulointerstitial microscopic fields (0.25 mm^2^) per section, and combined using the following percentage scores: 0 = 0 to 5%; 1 = > 5 - 20%; 2 = > 20 to 50 %, and 3 = > 50% glomerular and tubulointerstitial structure stained.

### Cell culture experiments

The podocyte cell line (P1) used in this study was characterized in detail before.^24, 25^ Peripheral blood mononuclear cells (PBMCs) were isolated by ficoll-hypaque density gradient centrifugation, and cultured in 10 ml RPMI 1640 medium containing 10% heat-inactivated fetal bovine serum (FBS), 5% interleukin-2, and 5 mg/ml PHA (Cellular Products, Buffalo, NY, USA). Human monocytes-macrophages were isolated from HIV-infected PBMCs, based on their adherence to plastic culture dishes, and stained with an antibody against the CD-68 antigen (Dako, Glostrup, Denmark). For the attachment assays, cells were seeded on plastic culture dishes and exposed to hrFGF-2 or control buffer in triplicate wells for 72 hours. Then all cells were fixed, stained with Diff Quick (Baxter Healthcare, Leesburg, VA, USA), and counted under the microscope with a 20X magnification lens as reported before.^21^ For the syncytia formation assays, HIV-infected PBMCs were seeded on fibrin gels derived from plasma taken from children with a high viral load.^26^ The HIV in situ hybridization studies were done using an HIV-probe from Lofstrand Labs Ltc. (Gaithersburg, MD, USA) as described before.^26^

### Infection and survival assays

The infection assays were done as described before.^25^ Briefly, the plasmids for HIV-1 infectious clone pNL4-3; pNL4-3.Luc.E-R- and GHOST(3) CXCR4+/CCR5+ cells were obtained from the National Institutes of Health AIDS Research and Reference Reagent Program (Bethesda, MD, USA.) pNL4-3Luc was constructed by excising the DNA fragment with BamH I and Xho I containing the luciferase gene coding sequence from the pNL4-3.Luc.E-R-(pNL43Luc-env) vector and inserting it into the same sites of pNL4-3. Recombinant HIV-1 was prepared in 293T cells transfected with the pNL4-3Luc vectors using the Lipofectamine 2000 kit from Life Technologies (Carlsbad, CA, USA) following the manufacturer’s protocol. Forty eight hours post-transfection, viruses present in the supernatants were collected by centrifugation at 500 x g for 10 min and filtered through a 0.45-μm-pore-size filter. For the luciferase assay, cells were grown in DMEM supplemented with 10% FBS and penicillin (100 U/ml)-streptomycin (100 µg/ml), all from Gibco BRL, (Gaithersburg, MD,USA). Cells (2 × 10^4^ /well) were seeded on 24 well culture dishes, treated with the corresponding concentration of hr-FGF-2 and exposed to HIV/NL4-3-Luc or HIV/NL4-3-LucΔenv. Forty eight hours later, the cells were lysed in 1% Triton X-100 buffer and processed for the luciferase assay as previously described.^25^ The survival assays were done in cells cultured under similar tissue culture conditions and exposed to hr-FGF-2 for 78 hours as described before. ^21^

### Real time qRT-PCR

Total RNA was purified from frozen cell pellets with TRIzol Reagent from Invitrogen (Waltham, Massachusetts, USA) as described before.^21^ cDNAs were synthesized from 2µg RNA with Quantabio’s qScript™ cDNA SuperMix (VWR, Radnor Pennsylvania, USA) following the manufacturer’s instructions. Quantitative PCR was performed with GoTaq® qPCR Master Mix (Promega, Madison, Wisconsin, USA) using 1/40 of the reverse transcription reaction. The primers and amplicon sizes were: *ApoL1*, forward 5’-AGGCCAGGAAAAGAAAGAGC, reverse 5’-TCTTCGGAGGACATTGAACC, 119bp (ref. PMID:<u>24169640</u>); *TNF*, forward 5’-CTCTTCTGCCTGCTGCACTTTG, reverse 5’-ATGGGCTACAGGCTTGTCACTC, 135bp; *Gapdh*, forward 5’-ACAGTCAGCCGCATCTTCTT, reverse 5’-GACAAGCTTCCCGTTCTCAG, 259bp (ref. PMID: <u>24578133</u>). The quantitative PCR program was as following: 95°C for 3 min, followed with 40 cycles of 95°C for 15 sec and 60°C for 1 min.

### Statistical Analysis

Data are reported as the mean ± SD or SEM for continuous variables that were normally distributed and as proportions for categorical variables. All data sets were tested for normality with the Shapiro-Wilk and Kolmogorov-Smirnov tests, in addition to residual and Q-Q plots. The median and interquartile range (IQR) was used for non-normally distributed variables. Differences between two groups were analyzed by paired or unpaired *t* test whenever appropriate. Intergroup comparisons were analyzed by one-way analysis of variance (ANOVA). Non-parametric data were compared using the Mann-Whitney test or the Kruskal-Wallis-test for multiple group comparisons. Correlation analyses were performed using the Spearman’s correlation coefficient test. P values <0.05 were considered significant. The odds ratio (95% CI) was estimated with Fisher’s exact test. The statistical analysis was done using the Prism 9 software (GraphPad Software Inc., La Jolla, CA).

## RESULTS

### Patients characteristics

Plasma and urine samples were collected in 84 children (≤ 12 years) who completed the follow up criteria discussed above between January 1994 and December 2000 at CNH. The demographic characteristics and clinical outcome of all children followed at CNH during that time was described before.^1^ In the study group, almost all children were of African descent (96%), and ranged in age from six weeks to 12 years (median [IQR] = 6 years [4-8]) with an approximately equal male to female ratio. They acquired HIV-1 through vertical transmission (95%) or blood transfusions (5%). Approximately 15% (13/84) developed HIV-CKDs, and 85% of them were of African descent. There were no significant age differences between those who developed or not HIV-CKDs.

### Plasma and urinary FGF-2 levels

Excluding all patients with HIV-CKDs, children with a high viral load showed higher plasma levels of FGF-2, when compared to those with a low viral load (Figure 1A). The Spearman rank correlation test showed a significant association between high viral loads and high FGF-2 plasma levels (Figure 1B), and between high plasma FGF-2 levels and proteinuria measured by dipstick in urine samples collected at the same time (Figure 1D). When all children with HIV-CKDs were excluded, the urinary FGF-2 levels (Figure 1C) and the Spearman rank correlation test (r = 0.1408, 95% CI −0.08 to 0.36; p = 0.228) were not significantly different between those with high and low viral loads. However, high urinary FGF-2 levels were detected in all children with HIV-CKDs (Figure 1C). Children with HIV-CKDs continued to show elevated FGF-2 plasma levels one year after the first measurement (median-[IQR] = 42 [32-46] pg/ml; 95% CI 32-47). Overall, children showing high plasma and urine FGF-2 levels (> 2 SD above the mean) had 73-fold increased odds of developing HIV-CKDs (OR 73.00; 95% CI 9.53-791.3; p < 0.0001) when compared to those showing low plasma and urine levels.

**Figure 1.**
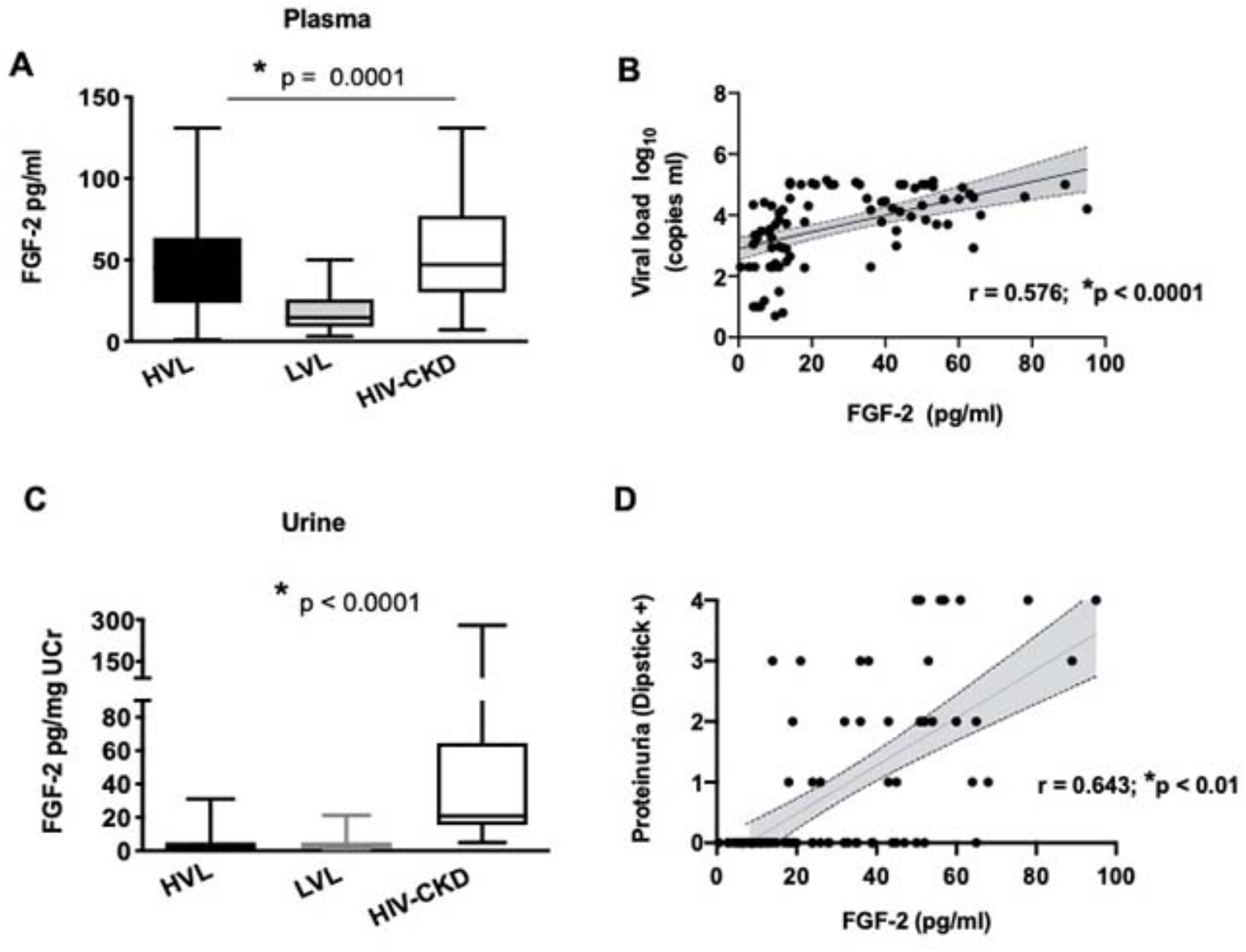
Association between FGF-2 plasma levels, viral load, and HIV-CKDs. Panel A shows the plasma levels of FGF-2 in children with high (HVL) and low (LVL) HIV-RNA viral loads, in comparison with those with HIV-chronic kidney diseases (HIV-CKD). HVL was defined as having at least three consecutive values > 5,000 or 3.7 log_10_ HIV-RNA copies/ml within 6-12 months prior to the FGF-2 measurement. Panel B shows the results of the Spearman rank correlation test done to determine the association between a high viral load and the FGF-2 plasma levels. Panel C compares the urine FGF-2 levels in children with high (HVL) and low (LVL) viral loads, excluding all children with HIV-CKD. Panel D shows the results of the Spearman correlation test done to define the association between the FGF-2 plasma levels and proteinuria. * P < 0.05 was considered statistically significant by one-way analysis of variance, Kruskal Wallis test.

### FGF-2 binding sites in the kidney of children with HIV-CKDs

The autoradiography binding studies done in frozen renal sections detected high affinity [^125^-I]-FGF-2 binding sites in renal glomeruli and vessels of all children (Figure 2A-B,E). However, children with HIV-CKDs showed an increased reservoir of low affinity FGF-2 binding sites in the renal interstitium, when compared to controls (Figures 2A-B). In agreement with previous studies, these bindings sites were displaced completely with 300 µg/ml heparin (Figure 2C-D).

**Figure 2.**
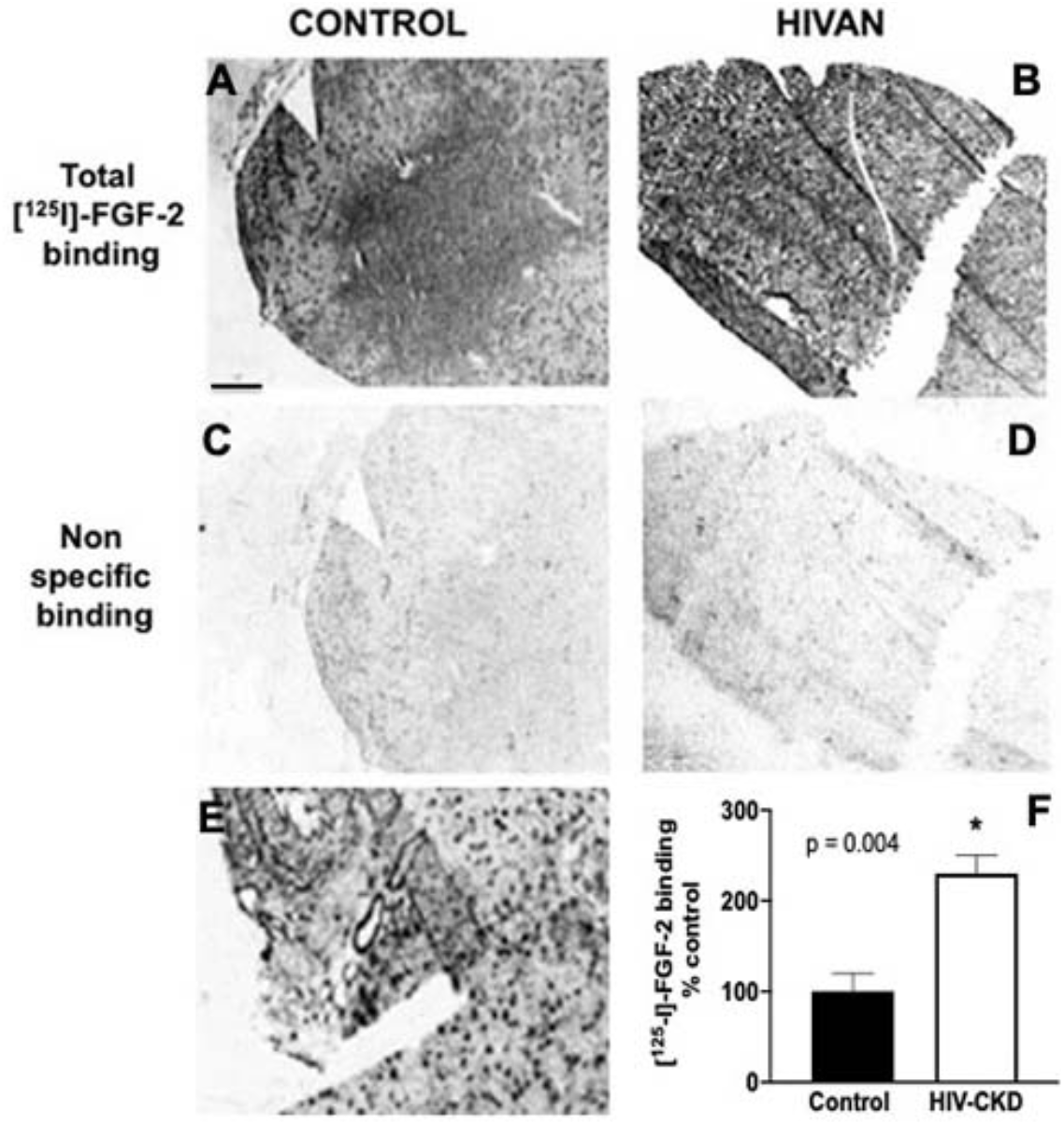
FGF-2 binding sites in the kidney of children with HIV-CKDs. Panels A and B show representative pictures of the total [^125^-I]FGF-2 binding detected by autoradiography in frozen renal sections derived from a control child (A) and a child with HIVAN (B). Panels C and D show representative pictures of the non-specific binding sites in similar sections. Panel E shows a picture of the high affinity [^125^I]-FGF-2 binding sites detected in renal glomeruli and vessels in a control child. Scale bars, A-E, 1mm. The graph in panel F shows the optical density [^125^-I]FGF-2 binding scores corresponding to children with and without HIV-CKDs. Error bars reflect the 95% CI. * p < 0.05 were considered statistically significant by the Mann-Whitney test (n = 3 per group).

The localization of FGF-2 binding sites in paraffin embedded renal sections was assessed with Alcian Blue, a cationic dye that at pH 1 binds predominately to the anionic sites of heparan sulfate proteoglycans (HSPG).^27^ As expected, the Alcian Blue staining was more intense in glomeruli, vessels, and the renal interstitium of children with HIV-CKDs (Figure 3A-B, D-E). Furthermore, by immunohistochemistry, FGF-2 was co-localized with the Alcian blue staining in the renal vessels (Figure 3G-H; Supplemental Figure 1), and accumulated in renal glomeruli of children with HIV-CKDs (Figure 3C,F, Supplemental Figure 1).

**Figure 3.**
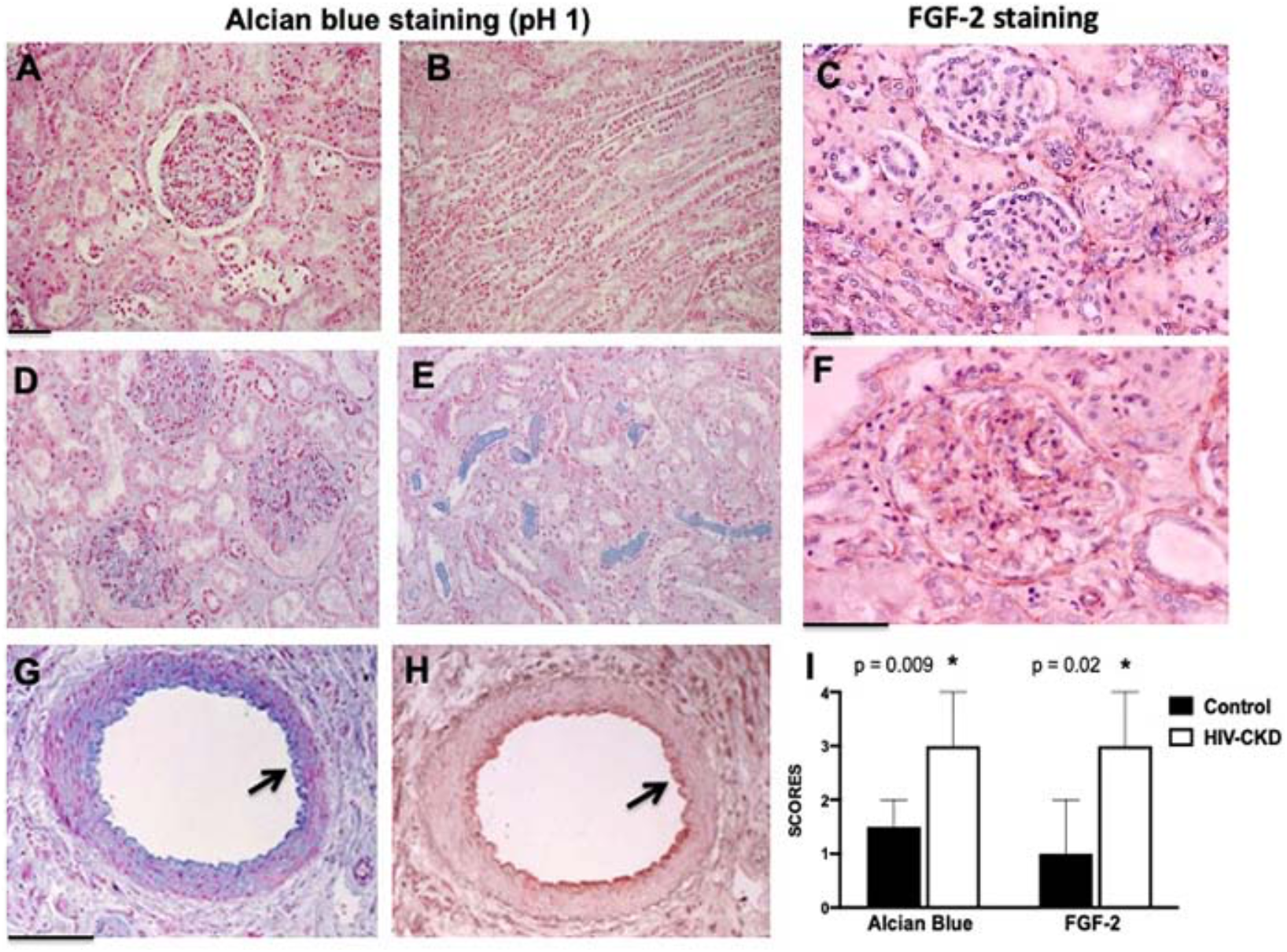
Increased Alcian blue and FGF-2 immunohistochemistry staining in renal sections from children with HIV-CKDs. Panels A and B show a representative Alcian blue staining (blue) in the renal cortex (A) and medulla (B) of a control child. Panel C shows a representative FGF-2 immunohistochemistry staining (red) in renal glomeruli from a control child. Panels D and E show a representative Alcian blue staining (blue) in the renal cortex (D) and medulla (E) of a child with HIVAN. Panel F shows a representative pictures of the FGF-2 immunohistochemistry staining (red) in a renal glomerulus from a child with HIVAN. Scale bars, A-F, 25 μm. Panels G and H, show a co-localization Alcian blue (blue) and FGF-2 staining (red) in renal vessels. The black arrow points to vascular endothelial cells. Scale bars, 50μm. The graph in panel I shows the Alcian blue and FGF-2 staining scores corresponding to control kidneys and those with HIV-CKD (n = 6 per group). Error bars reflect the 95% CI. P * < 0.05 was considered statistically significant by the Mann-Whitney test.

### Recruitment of circulating FGF-2 in the kidney of HIV-Tg_26_ with renal disease

To define whether more circulating FGF-2 was recruited in the kidney of HIV-Tg_26_ mice with renal disease, we injected [^125^ I]-FGF-2 intravenously to wild type (WT) and HIV-T_26_ mice. FGF-2 was rapidly cleared from the circulation in both groups of mice (*t*_1/2_ = 2 min), and accumulated mainly in the liver and kidney. More [^125^ I]-FGF-2 was detected however in the kidney of HIV-Tg_26_ mice with renal disease, and these mice showed a higher number of FGF-2 binding sites compared to WT mice (Figure 4A-C). The Alcian blue staining was also more intense in the kidney of HIV-Tg_26_ mice with renal disease (Figure 4D-E).

**Figure 4.**
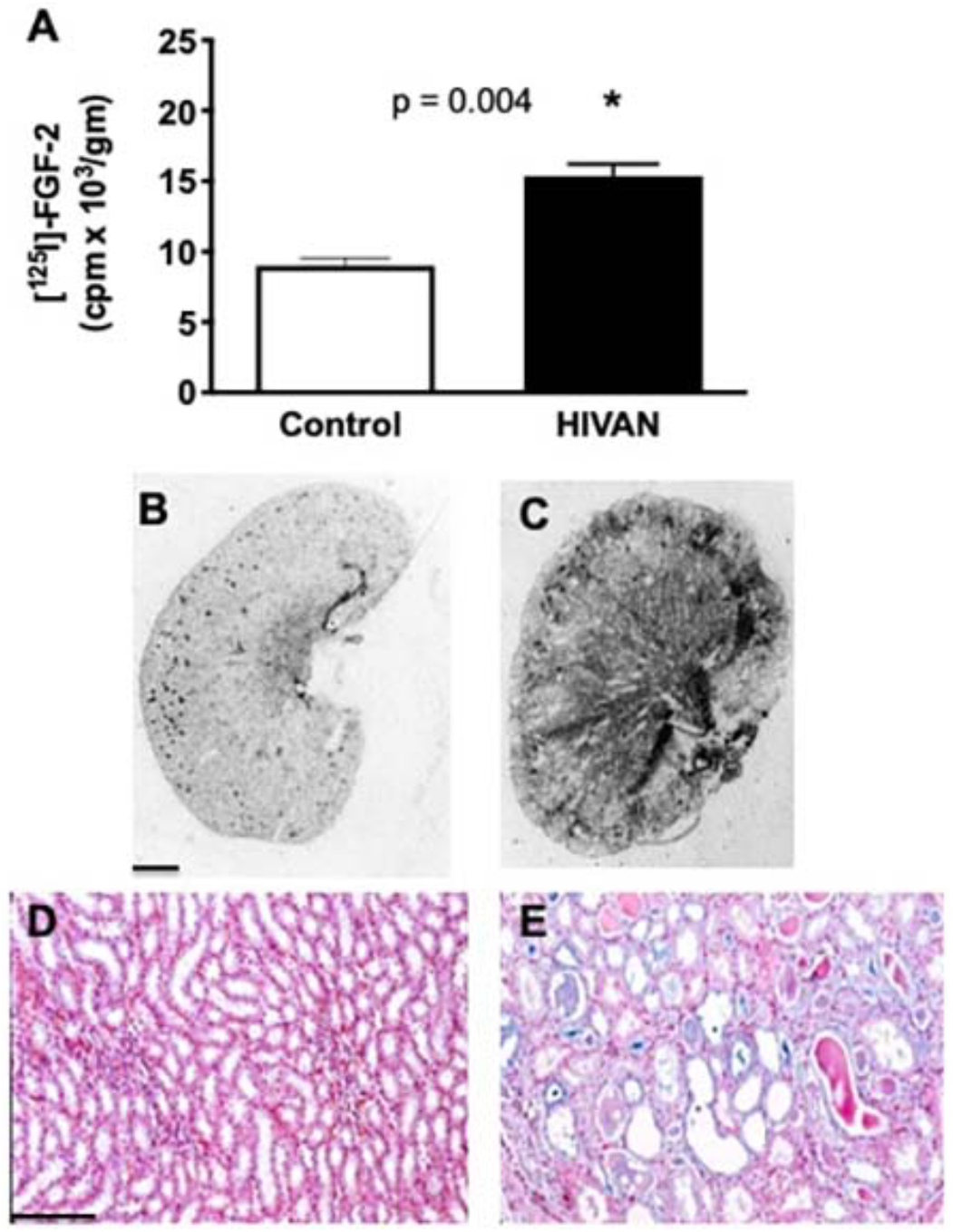
Recruitment of circulating FGF-2 in the kidney of HIV-Tg_26_ mice with renal disease. The graph in panel A shows how much [^125^-I] FGF-2 injected intravenously was detected in the kidney of control and HIV-Tg_26_ mice with renal disease. * P values < 0.05 were considered statistically significant by the Mann-Whitney test (n = 3 mice per group). Panels B and C show representative pictures of the [^125^-I]FGF-2 binding sites in the kidney of wild type and HIV-Tg_26_ mice with renal disease. Scale bars, 1.5 mm. Panels D and F show representative pictures of the Alcian blue staining in the renal medulla of wild type and HIV-Tg_26_ mice with renal disease. Scale bars, 100 μm.

### Circulating FGF-2 induced glomerular injury in HIV-Tg_26_

To determine whether circulating FGF-2 recruited in the kidney was capable of inducing glomerular injury, we injected hr-FGF-2 into young wild type and HIV-Tg_26_ FVB/N male mice without pre-existing proteinuria (10 µg/mouse/day x 7 days). By electron microscopy, we found that FGF-2 induced endothelial swelling, and mild fusion of the foot processes of podocytes in wild type mice (Figure 5A,C). FGF-2 also precipitated the collapse of glomerular in HIV-Tg_26_ mice (Figure 5D), mimicking the lesions seen in HIV-Tg_26_ mice that developed HIVAN spontaneously (Figures 5B). Renal glomeruli of wild type and HIV-Tg_26_ mice injected with FGF-2 showed more cells expressing PCNA, relative to wild type mice injected with PBS (Figure 6A-D), and these changes were associated with the development of proteinuria and renal injury (Figure 6E-F).

**Figure 5.**
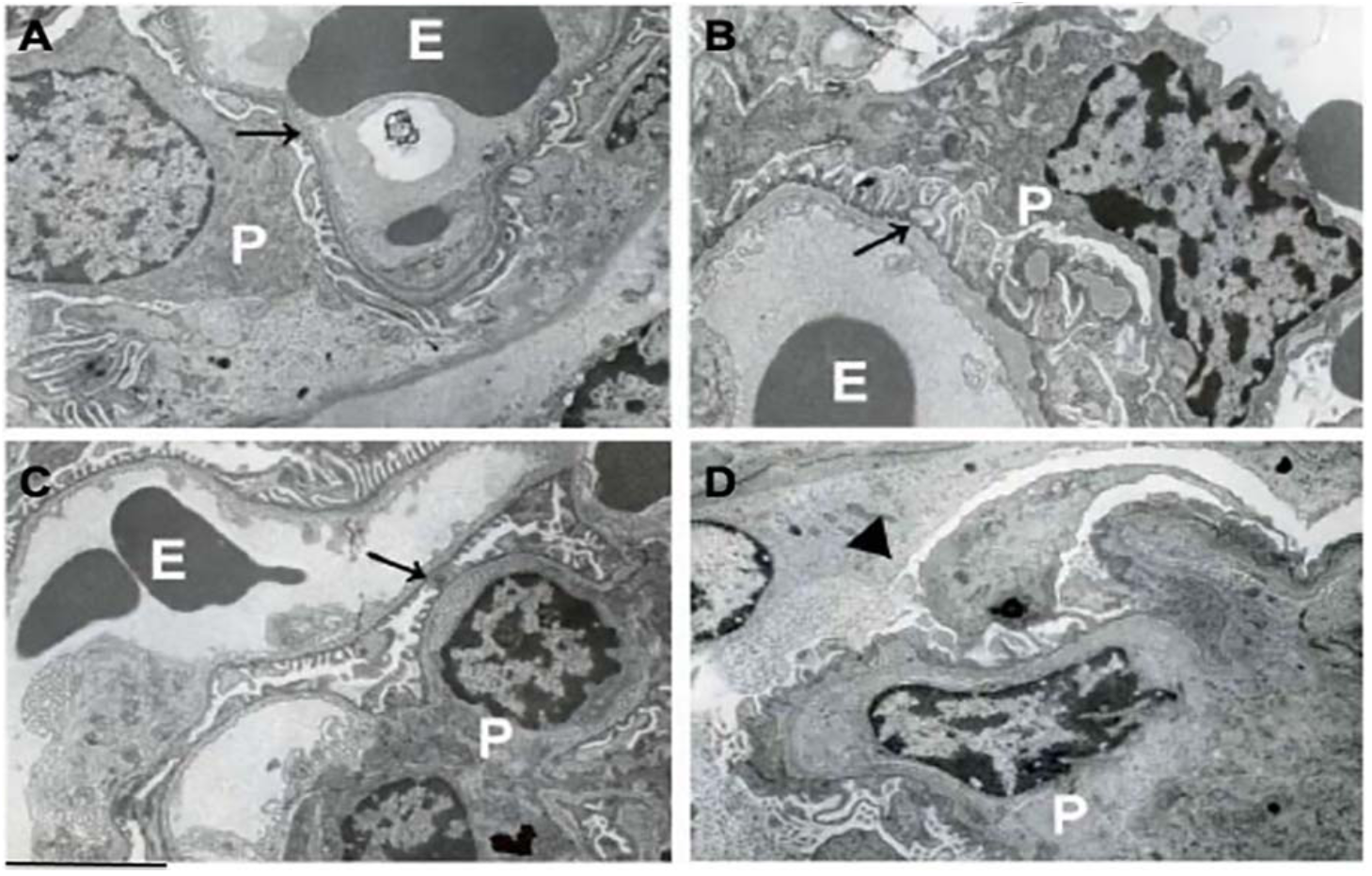
Circulating FGF-2 induced HIVAN-like lesions in HIV-Tg_26_ mice. The panels show representative electron microscopy pictures of renal glomeruli in mice injected intravenously with control PBS buffer or hrFGF-2 (10 µg/mouse/day x 7 days). Panel A shows a representative picture of normal glomerular endothelial cells and podocytes (black arrow) in wild type mice injected with PBS (E = erythrocytes; P = podocytes). Panel B shows foot processes effacement and enlarged podocytes (black arrow) in an HIV-Tg_26_ mouse which developed HIVAN spontaneously. Panel C shows a representative picture of the glomerular endothelial swelling (black arrow) and mild fusion of the podocyte foot processes in a wild type mouse injected with hr-FGF-2. Panel D, shows a representative picture of a collapsed glomerular capillary (arrowhead), swollen endothelial cells, and foot process effacement in an HIV-Tg_26_ mouse injected with FGF-2. Scale bars, A-D 2 µm.

**Figure 6.**
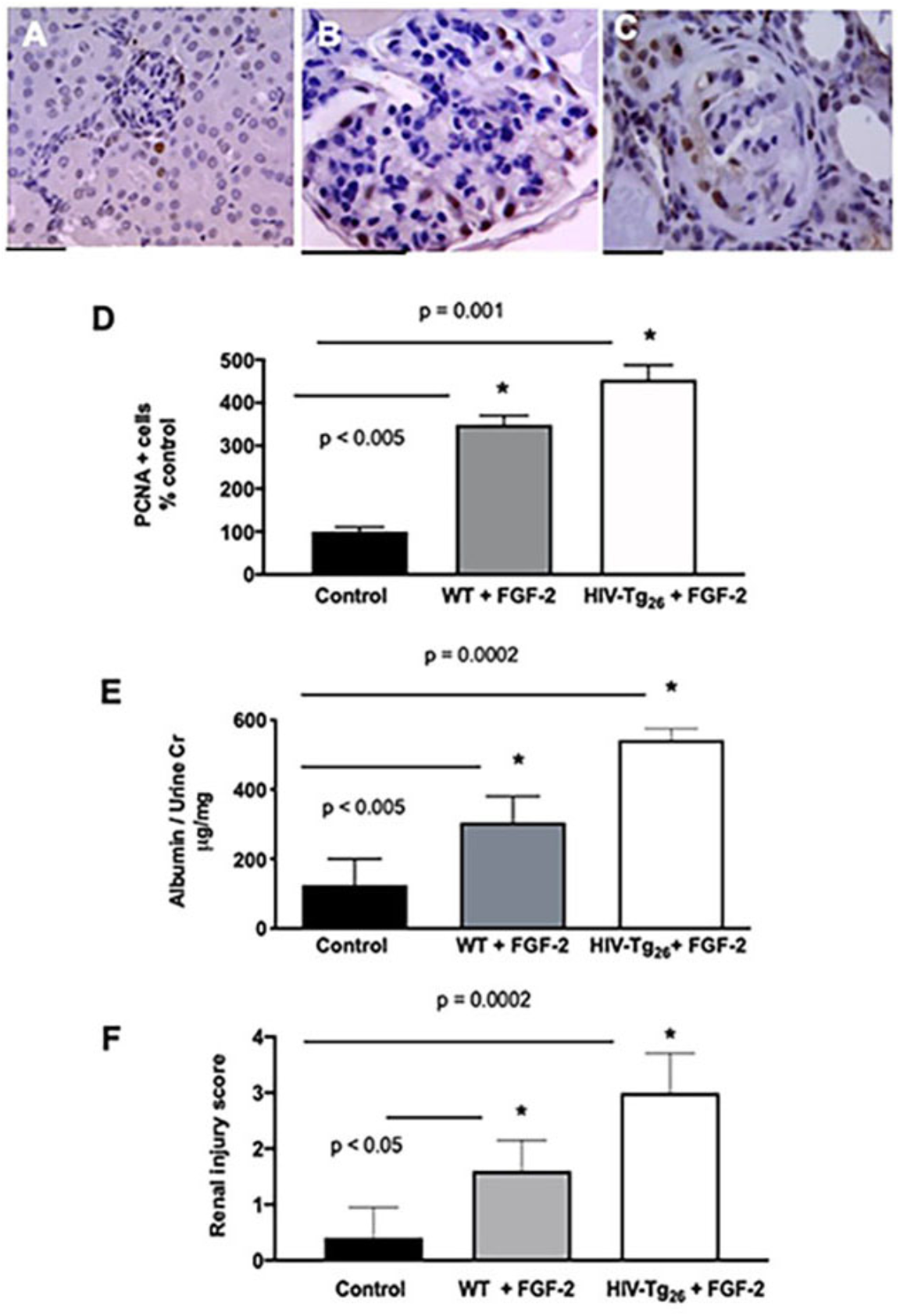
FGF-2 induced the expression of the proliferating nuclear cell antigen (PCNA), and induced proteinuria and renal injury in wild type and HIV-Tg_26_ mice. Panels A-C show a representative pictures of the PCNA staining (brown) in renal glomeruli of wild type mice injected with either PBS (A) or hr-FGF-2 (B), and in HIV-Tg_26_ mice (C) injected with hr-FGF-2. Scale bars, 25 μm. Panel D shows the quantitative scores for PCNA in five mice per group. Panel E shows changes in the albumin/urinary creatinine ratio in five mice per group. Panel F shows the renal injury scores in five mice per group. Error bars reflect the 95% CI. *P values < 0.05 were considered statistically significant by one-way analysis of variance, Kruskal Wallis test.

### FGF-2 increased the attachment, syncytia formation, and survival of HIV-infected cells

Since FGF-2 can increase the recruitment of inflammatory cells, we explored how FGF-2 affected the attachment and syncytia formation of cultured monocytes-macrophages derived from children with HIV-CKDs. Briefly, we found that both processes were enhanced by FGF-2 (Figure 7). In addition, we found that FGF-2 increased the survival of both podocytes and GHOST(3)-CXCR4+/CCR5+ cells infected with HIV-1 (Figure 8). Finally, we explored how FGF-2 affected the expression of Tumor Necrosis Factors-*α* (TNF-*α*) and APOL1 in podocytes, given that both molecules can either facilitate the infection and/or decrease the survival of these cells. By real time qRT-PCR, we found a minor but significant reduction in the steady state mRNA levels of both molecules in podocytes exposed to FGF-2 (Figure 9).

**Figure 7.**
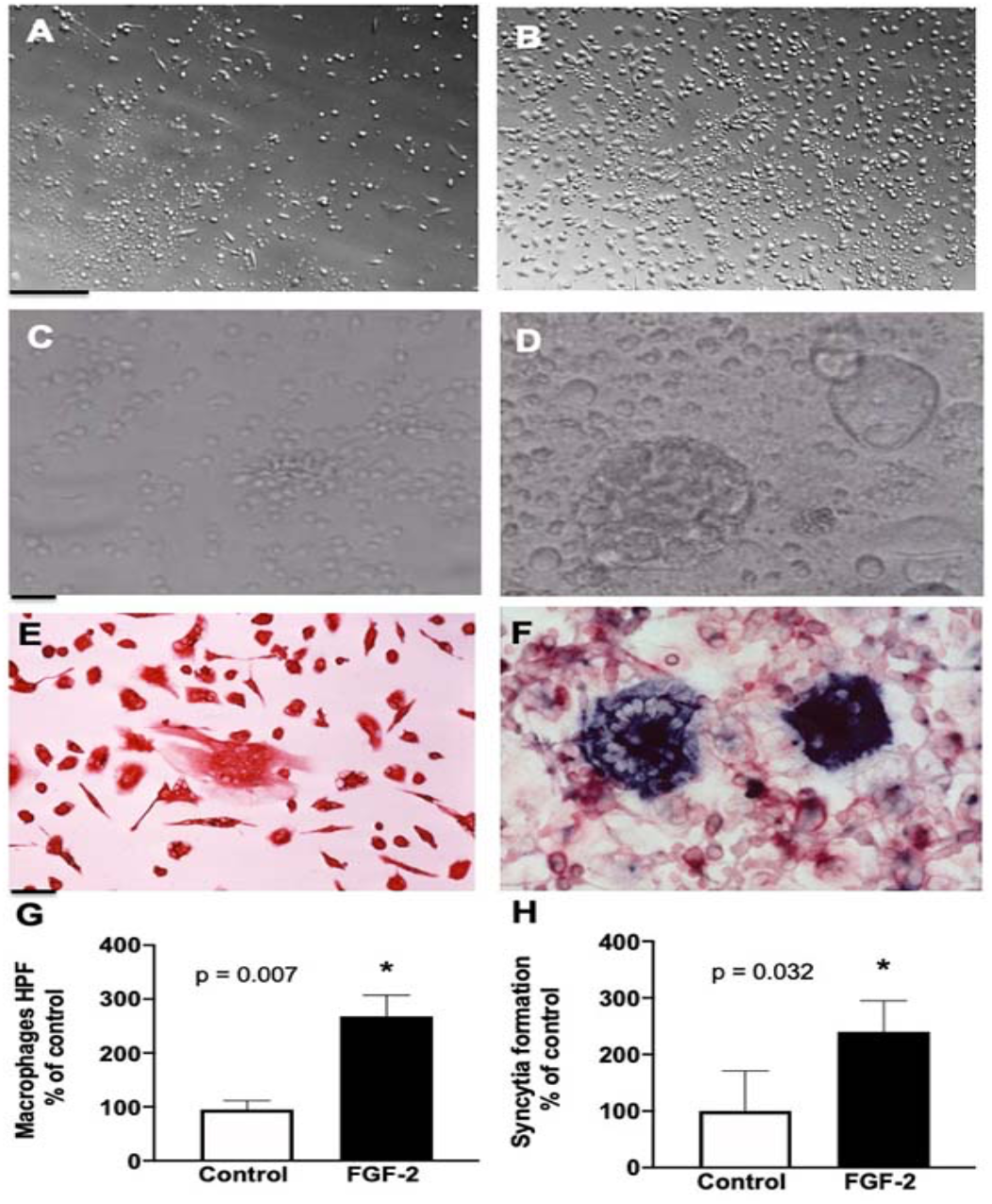
FGF-2 increased the attachment and induced syncytia formation in cultured human monocyte-macrophages. Panels A and B, show representative pictures of human monocytes-macrophages treated with either PBS (A) or hr-FGF-2 (20 ng/ml) for three days (B). Scale bars, 100 μm. Panels C and D show representative pictures of syncytia formation in HIV-infected peripheral blood mononuclear cells seeded on fibrin gels derived from plasma taken from a child with a high viral load, and exposed for five days to either PBS (C) or FGF-2 (20 ng/ml). Scale bars, 50μm. Panel E shows cultured human macrophages stained red with an antibody against the CD-68 antigen. Panel F shows a co-localized HIV-RNA staining (blue color by in situ hybridization) and CD-68 staining (red color by immunohistochemistry) in HIV-infected mononuclear cells. Scale bars, 25μm. The graphs, in panels G and H, show the quantitative scores corresponding to the attachment of monocyte-macrophage and their ability to form syncytia (n = 6 samples per group). The Error bars in the graphs reflect the 95% CI. *P < 0.05 were considered statistically significant by the Mann-Whitney test.

**Figure 8.**
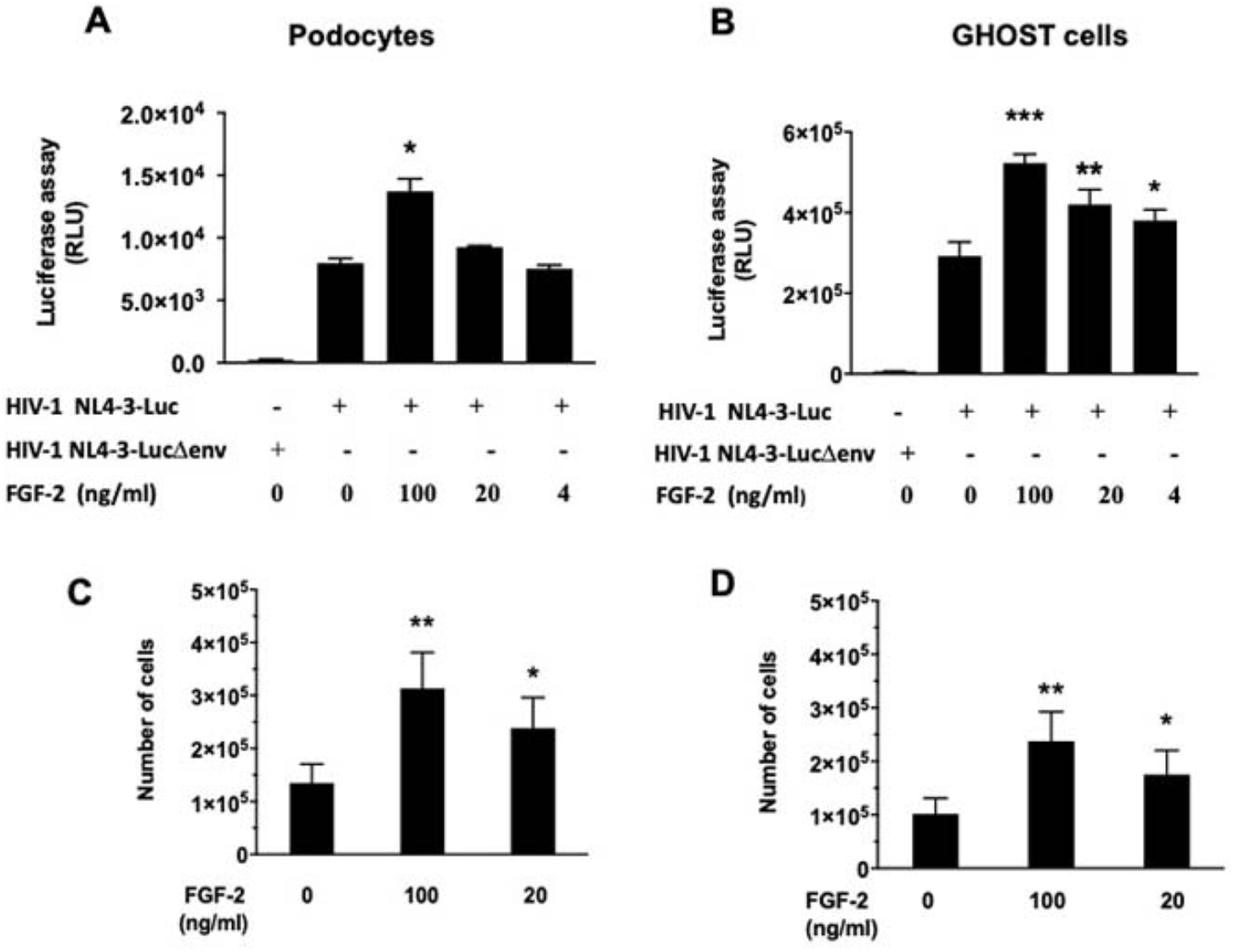
FGF-2 increased the survival of podocytes and GHOST(3) CXCR4+CCR-5+ cells infected with HIV-1. Panels A and B show podocytes (P1) and GHOST(3) CXCR4+ CCR5+ cells (human osteo sarcoma cells stably transduced with a CD4 retroviral vector and expressing the HIV-coreceptors CXCR4 and CCR5) infected with HIV-1 NL4-3-Luc and exposed to hr-FGF-2 for 48 hours. Panels C and D, show the survival studies done in these cells subjected to similar tissue culture conditions, as described in the Methods section. Error bars reflect mean ± SD. P values < 0.05; ** < 0.01; *** 0.001 were considered statistically significant.

**Figure 9.**
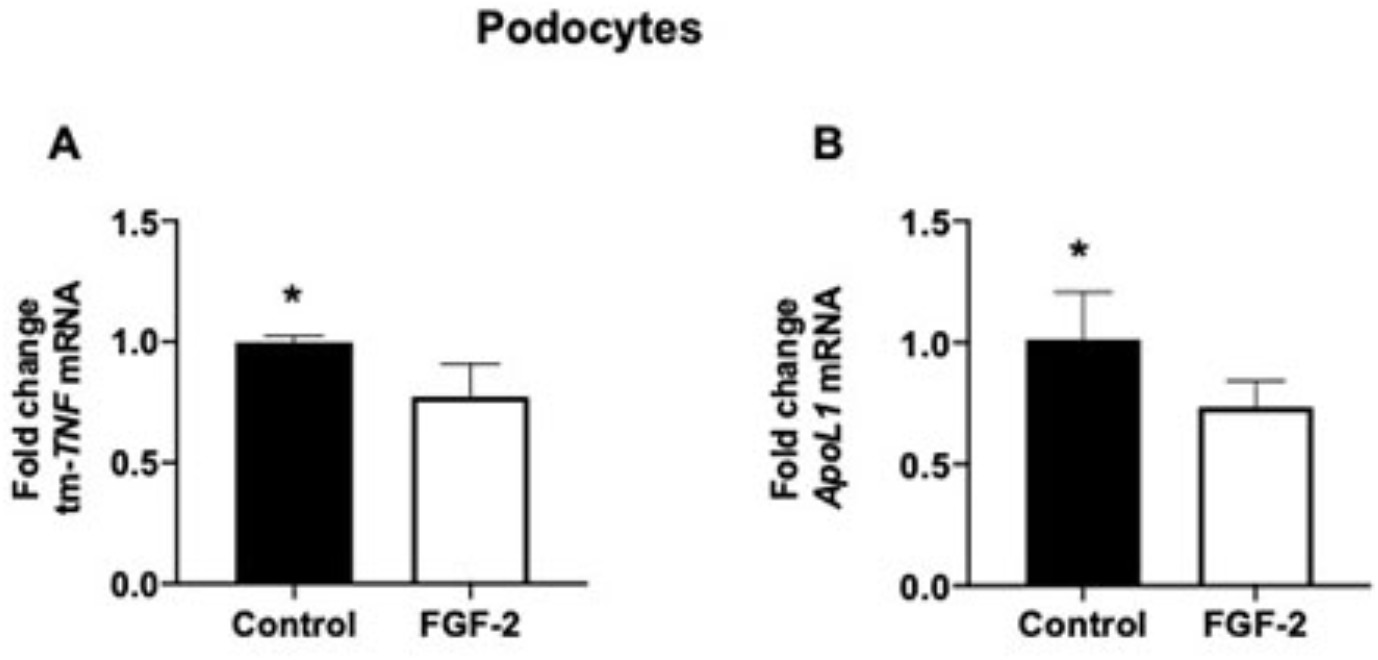
FGF-2 decreased the steady state mRNA levels of TNF-α and APOL1 in cultured podocytes. One x 10^5^ podocytes (P1) were seeded in triplicate and exposed to control PBS buffer or rh-FGF-2 (20 ng/ml) for 48 hours. The Real Time qRT-PCR analysis for TNF-*α* and APOL1 was done in RNA extracted from podocytes as described in the Methods section. Error bars reflect 95% CI. * P values < 0.05 were considered statistically significant by the Mann-Whitney test.

## DISCUSSION

In this study, we showed that high plasma levels of FGF-2 in young children living with HIV were associated with a high viral load and HIV-CKDs. We also found that children with HIV-CKDs had a large reservoir of kidney FGF-2 low affinity binding sites that can facilitate the recruitment of circulating FGF-2. In addition, we reported that circulating FGF-2 induced HIVAN-like lesions in HIV-Tg_26_ mice, and increased the survival of cultured mononuclear cells and podocytes infected with HIV-1. Overall, children with both high plasma and urine FGF-2 levels, had 73-fold increased odds of developing HIV-CKD compared to those with lower plasma and urine FGF-2 levels.

FGF-2 is a heparin binding growth factor that lacks a classic signal peptide for secretion, and it is stored bound to HSPG in the subendothelial matrix and basement membranes.^28, 29^ Therefore, very low FGF-2 levels are detected in the circulation of healthy children. However, in young children living with HIV, FGF-2 can be released through non-conventional pathways, including endothelial injury and inflammatory cells.^30-32^ Nonetheless, the role of circulating FGF-2 in the pathogenesis of childhood HIV-CKDs is unknown. To address this issue, we measured the plasma and urine levels of FGF-2 in young children living with HIV between 1994 and 2000, given the high prevalence of HIV-CKDs in young children at that time. Only children who received standard medical care and completed at least three consecutive years of medical follow up at CNH qualified as participants. Thus, the viral loads and FGF-2 plasma levels reported here, reflect the clinical status of children living in the US during the pre-modern ART era. Our definition of a high viral load, was also based on the standard of care at that time, since viral loads > 5,000 HIV-RNA copies/ml (> 3.7 log_10_) were associated with worse clinical outcomes.^17^ In contrast, no significant differences in clinical outcomes were noted at that time, between people with viral loads ranging from 5,000 to 500 (2.7 log_10_) HIV-RNA copies/ml.^17^ However, our findings are relevant for the large number of children living with HIV that are not receiving appropriate ART today, and in agreement with the high serum levels of FGF-2 detected in adults living with HIV.^9^ In addition, they are consistent with the notion that many cytokines released by HIV-infected cells (interferon *γ* IL-1*β*, and TNF-*α*) can facilitate the release of FGF-2 by non-conventional mechanisms.^32^

FGF-2 plays important roles during renal development^12, 33^, regeneration^34, 35^ and maintaining the blood pressure^36^, and vascular tone.^37, 38^ Previous studies showed that FGF-2 acting through its high and low affinity receptors can induce the proliferation of almost all renal cells types ^13-16, 39^ and participates in the pathogenesis of many human renal diseases.^40-42^ In addition, the plasma and urine FGF-2 levels in children have been considered reliable candidate biomarkers to predict the outcome of reflux nephropathy^43^, renal cysts^44, 45^, the Hemolytic Uremic Syndrome^46^, and other renal and vascular complications in critically ill children.^47, 48^ Furthermore, children with HIV-CKDs showed high expression levels of a Fibroblast Binding protein (FBP-1) that increases the renal mitogenic activity of FGF-2. ^20, 39^ Taken together, all these data suggest that the renal accumulation of FGF-2 may accelerate the progression of childhood HIV-CKDs.

Under normal conditions FGF-2 is rapidly cleared from the circulation by vascular endothelial cells and sequestered in the subendothelial matrix^49^, where it remains silent protected from proteolytic degradation.^50^ Our findings are in agreement with this notion, although circulating FGF-2 induced mild glomerular injury and transient proteinuria in wild type mice. This observation, is in agreement with the kidney lesions seen in rats and monkeys injected with FGF-2 for months, which cause podocyte hypertrophy and focal glomerular sclerosis^10, 11^, as seen in children with HIVAN. The exact mechanism by which circulating FGF-2 induces glomerular injury, is unclear. It has been postulated that podocytes are terminally differentiated cells, and when they are forced to enter the mitotic cycle, they cannot undergo cytokinesis and detach or die.^51^ In agreement with this hypothesis, we found that FGF-2 increased the expression of PCNA in wild type podocytes. We also showed that FGF-2 induced HIVAN-like lesion and gross proteinuria in HIV-Tg_26_ mice. The latter findings suggest that podocytes that are primed or injured by HIV-genes or cytokines may be more sensitive to FGF-2. This notion is consistent with previous reports showing that HIV-Tat acts in synergy with FGF-2 to disrupt the cytoskeletal structure of podocytes and glomerular endothelial cells.^24, 52^

As discussed above, HSPG can act as FGF-2 low-affinity receptors.^53^ The interaction between HSPG and FGF-2 promotes the dimerization of the FGF high-affinity receptors increasing their phosphorylation and activity.^53^ In this manner, HSPG can serve as a sink to concentrate, stabilize, and regulate the activity of FGF-2 in the kidney. Syndecan, perlecan, and glypican, are key HSPG involved in this process.^54, 55^ For example, mutations that affect the binding of FGF-2 to glypican 5 can impair its ability to induce proteinuria in mice with hyperglycemia or treated with puromycin.^56, 57^ Given the limited number of renal sections available from children with HIV-CKDs, we used the cationic dye Alcian blue, which at pH1 binds to all HSPG, to determine the potential localization of the FGF-2 low affinity bindings sites in paraffin embedded renal sections. As expected, all renal structures that stained more intensely with Alcian blue, also showed increased FGF-2 binding affinity and more significant FGF-2 staining. Thus, a novel finding of our study is the identification of a simple and cost-effective method to assess the localization of FGF-2 binding sites. This staining may be useful in clinical practice to identify childhood renal diseases at higher risk of progression by accumulating FGF-2.

Previous studies showed that FGF-2 increases the recruitment of inflammatory cells.^58, 59^ Lymphocytes and macrophages release FGF-2^60^, and two molecules that facilitate the migration of lymphocytes and macrophages, L-selectin and monocyte chemoattractant protein-1 (MCP-1), also bind to kidney HSPG.^61^ A key step in the pathogenesis of HIV-CKDs is the renal recruitment of HIV+ mononuclear cells, which facilitates the infection of epithelial cells through cell-to-cell transfer mechanisms.^26^ HSPG are involved in this process by facilitating the attachment and entry of HIV-1 via endocytosis.^25^ Therefore, we carried out additional studies to determine how FGF-2 affected the infection of inflammatory cells and podocytes. We found that FGF-2 increased the attachment, survival, and syncytia formation of HIV+ macrophages cultured from children with HIV-CKDs. In addition, FGF-2 increased the survival of HIV-infected podocytes and decreased the steady state levels of TNF-*α* or APOL1 mRNA, two molecules that either facilitate the infection^25^ and/or decrease the survival of these cells.^62^

In summary, a high viral load facilitates the release of FGF-2 through different mechanisms. In addition, an up-regulated expression of the FGF-2 low affinity binding sites (HSPG) in the kidney, facilitates the renal recruitment of circulating FGF-2. Finally, by increasing the attachment and survival of HIV-infected cells, FGF-2 appears to perpetuate the renal injury process. Taken together, these findings support the novel concept that sustained high plasma FGF-2 levels may constitute an independent risk factor for the development of HIV-CKDs in children.

## Data Availability

All data presented have been collected at CNH and approved by the IRB.

## Acknowledgment

We thank Dr. Tamara Rakusan and all physicians and nurses that contributed to the medical care of these children during the study period. This study was supported by National Institutes of Health awards: R01 DK-103564; R01 DK-108368; R01 DK-115968; R01 DK-04941, and HL −102497.

