## Supplemental Figure 1 for "Association of circulating Fibroblast Growth Factor-2 with progression of HIV-chronic kidney diseases in children"

SUPPLEMENTARY MATERIAL

**Supplemental Figure 1.** *FGF-2 and Alcian blue staining in renal glomeruli and vessels.* Panel A shows renal glomeruli from a child with HIV-HUS exposed to the control primary antibody (non-specific IG) as described the Methods section. Panel B shows renal glomeruli from the same child with HIV-HUS stained with the FGF-2 antibody (red). Panels C and D show renal vessels stained with Alcian blue (C) or the FGF-2 antibody (red). Scale bars, A-D, 25 $\mu$m. (PDF file)

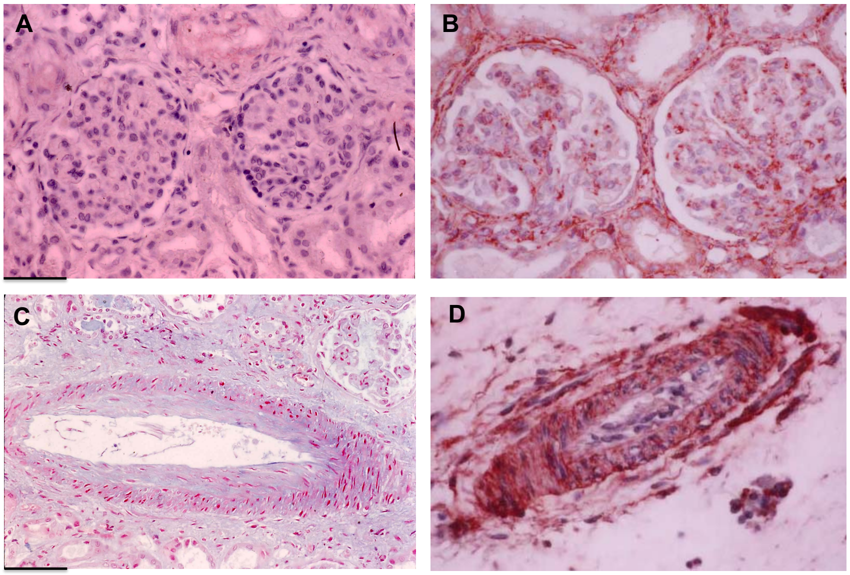
